# A hybrid care intervention for high-risk patients with chronic obstructive respiratory diseases: bridging the gap between clinical trials and real-world practice

**DOI:** 10.1101/2025.09.11.25335555

**Authors:** Alba Gómez-López, Núria Sánchez-Ruano, Marta Sorribes, Rubèn Gonzalez-Colom, Marina Paredes, J S You, MJ Gordillo, Emili Vela, Gerard Carot-Sans, Alba Jimenez, Xavier Michelena, Ramon Farre, Josep Roca, Isaac Cano, Alicia Aguado, Jose Fermoso, Néstor Soler, Ebymar Arismendi

**Author notes:** Corresponding author: Rubèn González-Colom, C/ Rosselló, 149, 08036, Barcelona, Spain. Equal contribution as senior authors (Néstor Soler, Ebymar Arismendi).

## Abstract

**Background:** Community-based management of exacerbations in high-risk patients with chronic obstructive respiratory diagnoses remains a major challenge due to patients’ heterogeneities, comorbidities and symptoms-based assessment of the episodes. Hybrid care interventions, combining digital tools with in-person, patient-centered care, have demonstrated efficacy in reducing unplanned hospitalizations. However, their effectiveness in real-world settings is less well established.

**Objectives:** To conduct a co-design process aiming to: (1) identify target candidates; and (2) adapt the implementation of a hybrid care intervention to routine clinical practice.

**Methods:** In the Integrated Health District of Barcelona-Esquerra (520 k citizens), four Plan-Do-Study-Act (PDSA) co-design cycles, each lasting six months, were conducted using a mixed-methods approach during the two-year follow-up (2024–2025) of a cohort of 205 high-risk patients with chronic obstructive pulmonary disease (COPD) and co-morbidities or severe asthma.

**Results:** By the end of PDSA-3, we profiled the target candidates for the hybrid care intervention and refined its main components: i) health risk assessment, ii) digital support with an adaptive case management approach, and iii) nurse-led in person care. Home-based, patient’s self-administered Oscillometry was feasible and acceptable for the objective assessment and management of exacerbations. Adapting the implementation of the hybrid care intervention to local clinical workflows was identified as a priority to enable its sustainable adoption.

**Conclusions:** The adaptive case management approach for personalized hybrid care at community level is suitable and shows acceptability by the different stakeholders. Its sustainable adoption and scalability in the real-world setting still requires implementation tailoring and demonstration of healthcare value generation.

**Trial registration number:** NCT06421402.

## INTRODUCTION

Patients with chronic obstructive pulmonary disease (COPD) or severe asthma frequently experience unplanned hospitalizations, partly explained by suboptimal management of multifactorial acute episodes at the community level[1–3]. These events deteriorate health-related quality of life, worsen prognosis and disproportionately consume healthcare resources[4]. A substantial proportion of these admissions can be considered potentially avoidable hospitalizations, highlighting the urgent need for effective preventive strategies at the community level[5]. While consensus guidelines exist for inpatient management of severe exacerbations in these patients[6,7], effective community-based strategies remain an urgent priority[4,8].

Evidence indicates efficacy of early detection and personalized community-based care to reduce hospitalization rates, improve clinical outcomes, and enhance quality of life, while simultaneously alleviating strain on healthcare systems, addressing a critical unmet need in global health policy[9– 12]. However, a persistent gap remains between the outcomes of randomized controlled trials (RCTs) and the real-world effectiveness of the approach[13]. This is partly due to current definitions of exacerbation rely broadly on symptomatic changes. Despite the application of accepted clinical guidelines[1–3], the inherent heterogeneities in patients with diverse multimorbidity profiles, among other factors, add complexity to the management of these patients[14–16].

The current study builds-up on several sources of evidence[12,13,17,18] indicating that personalized nurse-led hybrid care interventions with a patient-centred approach, in which in-person care and digital support are combined, can facilitate early community-based management of exacerbations to effectively prevent emergency room consultations and unplanned hospitalizations.

To address this efficacy-effectiveness gap, we initiated a two-year follow-up of a cohort of 205 patients with chronic obstructive respiratory disorders showing high risk for exacerbations[19]. In this cohort, we are piloting a nurse-led hybrid care intervention whose seminal characteristics have been recently described [20].

The study protocol of this hybrid care intervention includes the refinement of its different components conducted through a co-design process, involving all stakeholders, through four Plan-Do-Study-Act (PDSA) sequential cycles[20,21], each of them lasting six months.

At the end of the co-design process the hybrid-care intervention is expected to enhance routine management of these patients in two well-differentiated ongoing clinical programs: (1) Community-based management of high-risk chronic obstructive respiratory patients living in the Integrated Health District of Barcelona-Esquerra[22] (AISBE, 520 k citizens), and (2) Severe Asthma Unit at Hospital Clinic de Barcelona (HCB).

The underlying hypothesis is that a tailored implementation of the nurse-led hybrid care intervention facilitates its sustainable adoption and scalability in real-world settings, beyond the current study cohort. Therefore, the objectives of this paper are twofold: (1) to characterize the profiles of high-risk obstructive respiratory patients who are candidates, aiming at personalizing the hybrid care intervention, and (2) to describe the co-design process used to adapt this intervention for its implementation in routine clinical practice[20]. The ongoing co-design process is aligned with recent WHO’s recommendations[23] on the need for development and sustainable implementation of the digital health transformative potential.

## METHOD

### Study design and participants

We conducted a mixed-methods study[24] in the catchment area of HCB (AISBE, Barcelona, ES), reporting qualitative co-creation activities in parallel with quantitative profiling of participants’ clinical characteristics and digital literacy, in accordance with guidelines for mixed-methods studies. [24,25]

Between November 2023 and March 2025, we offered enrolment to all individuals visited in the context of two distinct clinical programs: (1) a community-based program for patients with chronic obstructive respiratory diseases (i.e. COPD, asthma, or bronchiectasis) and a high comorbidity burden, defined as a risk score above the 80^th^ percentile on the Adjusted Morbidity Groups (AMG) scale[26,27] (managed in AISBE), and (2) patients with severe asthma[28,29] managed at HCB’s Severe Asthma Clinic. All participants were independent for daily life activities. Patients enrolled in intermediate care, home care, or palliative care programs were excluded. Further details on the inclusion and exclusion criteria are listed in the study protocol [20].

### The co-design process

We implemented a co-design process to customize the hybrid care intervention, ensuring it aligns with local care pathways and supports its sustainable adoption. To achieve this, four sequential PDSA co-design cycles, each lasting six months, were conducted by a multidisciplinary research group[30] with the active contributions of all the stakeholders (**Appendix 1**). The outputs from the initial three co-design cycles were collectively evaluated by a dedicated focus group session with a fourteen leading professionals, mixing clinical staff (n=9), IT developers (n=1), data scientists (n=1) and managers (n=3) to consolidate findings and reach a final consensus on the structure of the integrated hybrid care intervention, and the implementation strategy, aiming at fostering sustainable adoption in routine practice.

### Patient profiling

Contextual data were collected to characterize the study population across multiple dimensions, including sociodemographic, clinical and functional characteristics, unhealthy lifestyle habits, multimorbidity burden[26,27,31], disabilities, social frailty, use of healthcare resources and digital literacy. Such comprehensive assessment of the study cohort aimed at identifying the candidates’ profiles for a personalized hybrid care intervention.

All data for patient profiling was gathered from three sources. At baseline, the nurse case manager instructed the patients, administered the different questionnaires and performed lung function testing, through two home visits of one hour duration each. Clinical information and therapies were gathered from the hospital care electronic records, with access to primary care information. Finally, we used the Catalan Health Surveillance System (CHSS)[32,33] for extracting historical data on resource utilization, multimorbidity and sociodemographic data. The CHSS centralizes data from primary and specialized care across the entire healthcare system.

### Clinical characterization

At baseline, the patients were clinically characterized using a comprehensive set of variables, including sociodemographic data, primary respiratory diagnosis, comorbidities, lung function using forced spirometry and Oscillometry, and healthcare resource utilization in the 12 months preceding enrolment (primary care visits, emergency department visits, hospital admissions, and total healthcare expenditure). Data on home-based respiratory therapies (oxygen therapy, non-invasive ventilation, and nebulized treatments) were also collected. Functional capacity was assessed using standardized patient-reported outcomes evaluating physical and mental health [34–36], as well as respiratory symptoms[37–41] and overall health status[20]. Additional variables included lifestyle risk factors (smoking, alcohol, physical activity, passive smoking), as well as housing characteristics[42] and living conditions.

### Digital literacy profiling

Digital literacy was assessed with a purpose-designed questionnaire comprising different items evaluating knowledge, usage, and confidence in digital skills relevant to the hybrid care model. Results were compared with a parallel classification into five levels of digital skills (None, Low, Medium with no improvement capacity, Medium with improvement capacity, High), based on the engagement and support needs observed by the nurse.

### Respiratory diagnoses profiling

To ensure diagnostic accuracy, the single respiratory diagnosis validated by the research team examining the patients’ electronic healthcare records was compared with the respiratory diagnoses obtained from the CHSS, including diagnostic information across all healthcare tiers.

### Statistical considerations

Continuous variables are reported as mean (SD) or median (IQR, defined as the 25^th^ and 75^th^ percentiles), as appropriate; categorical variables are presented as n (%). Between-group comparisons for continuous data used the Student’s-t test when distributional assumptions were met and the Wilcoxon rank-sum test otherwise. Categorical data were compared with Fisher’s exact test. All tests were two-sided, and p < 0.05 was considered statistically significant. All the analyses have been conducted using R version 4.1.1[43].

### Ethics approval and consent to participate

Ethical approval for the process evaluation was granted by the Ethical Committee for Human Research at the HCB on June 29, 2023 (HCB/2023/0126) and registered at ClinicalTrials.gov (NCT06421402). The patients participating in the research were required to provide and sign a written informed consent indicating the purpose of the study, the nature, use, and management of their data.

## RESULTS

### Characterization of the cohort

During the recruitment period, 205 patients were enrolled in the study: 152 (74%) from the community program (AISBE) and 53 (26%) from the severe asthma group (**Figure 1**). Of the 205 patients enrolled, 22 (10.7%) were excluded from analysis (19 withdrawals; 3 deaths), resulting in an analytic cohort of 183 participants. **Table 1** summarizes baseline clinical characteristics for (1) the overall analytic cohort (n = 183), (2) community-managed patients in the AISBE program (n = 135; 74%), and (3) patients followed at HCB’s Severe Asthma Unit (n = 48; 26%).

**TABLE 1.**
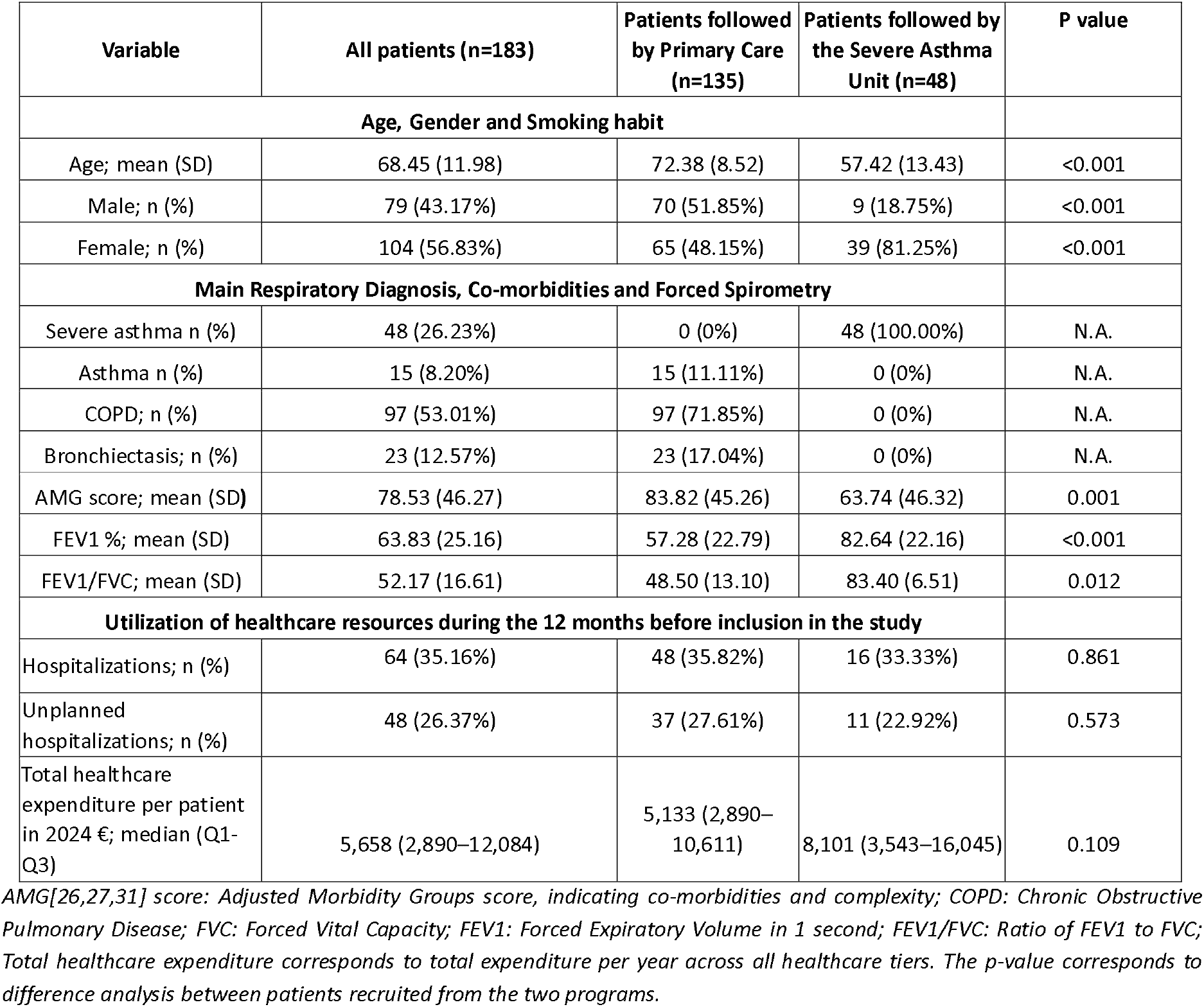
Main characteristics of the study groups.

**Figure 1.**
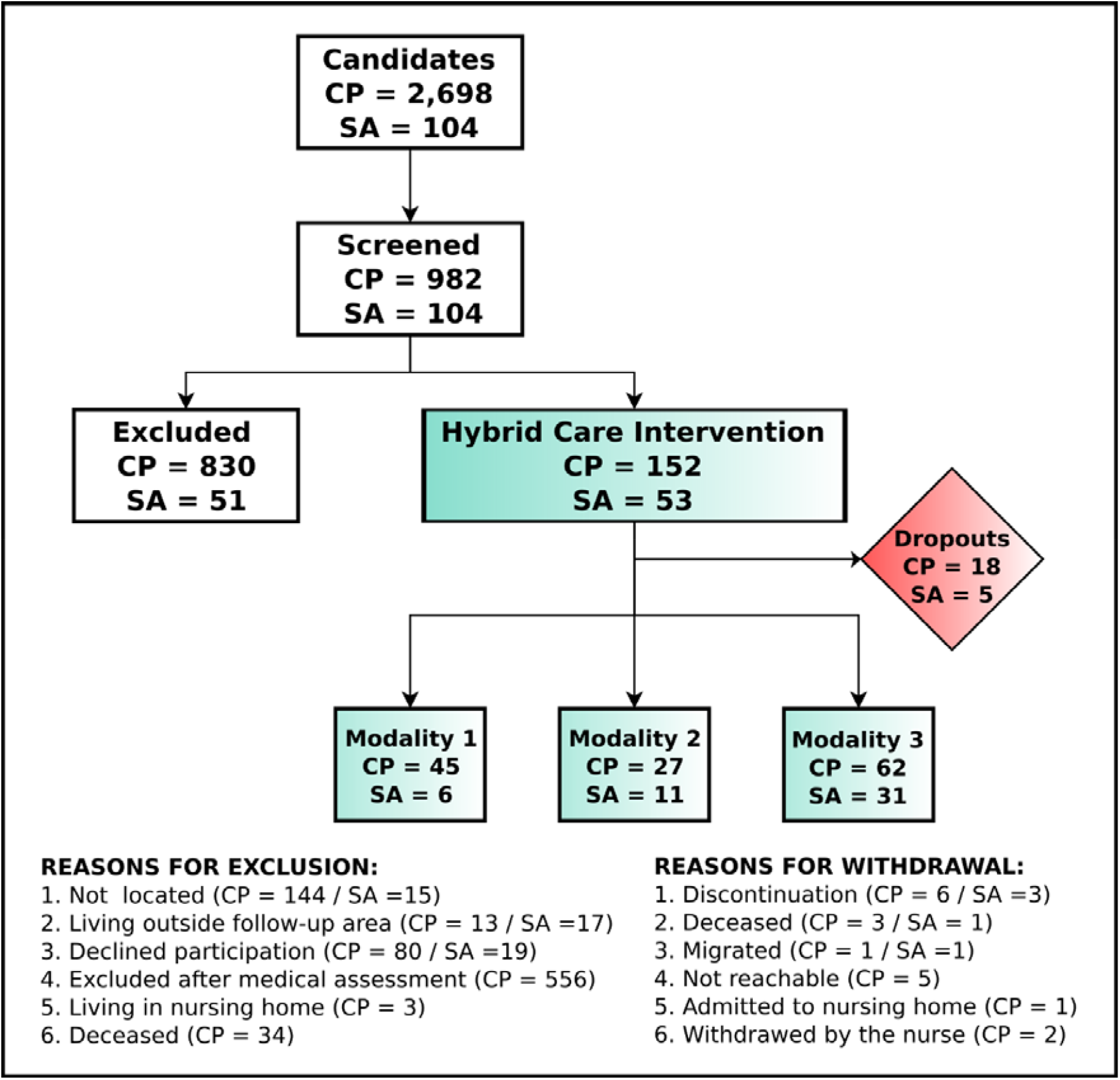
Study Flow Diagram. CP: Community-based program in AISBE; SA: Severe Asthma Unit program. Modalities 1-3 refer to the characteristics of the digital support to clinically stable patients during the follow-up: M1: Phone call–managed (28 %), M2: APP-managed (chat & PROMS) (21%) and M3:APP-managed (chat & PROMS) + HRV sensor (51%). All the modalities include indoor air quality (IAQ) monitoring with low-cost sensors [44].

Compared with patients from the Severe Asthma Unit, the community-managed patients with chronic obstructive respiratory diseases were older (72.4 vs 57.4 years; p<0.001), more often male (51.9% vs 18.8%; p<0.001), and had a higher comorbidity burden (AMG 83.8 vs 63.7; p=0.001). Lung function indicated greater airflow limitation in AISBE (FEV_1_% predicted 57.3% vs 82.6%; p<0.001; FEV_1_/FVC 48.5 vs 83.4; p=0.012). Healthcare utilization over the prior year was broadly similar between groups (all-cause hospitalization 35.8% vs 33.3%, p=0.861; unplanned 27.6% vs 22.9%, p=0.573). We found no significant differences in healthcare expenditure per patient between the AISBE group (€8,101) and the severe asthma group (€5,133) (p=0.109).

Regarding the validation of the respiratory diagnosis, 62% of the patients (n=114) showed coincidence between the respiratory diagnosis validated by the research team and the registry information. The regional registry (CHSS) contained two respiratory diagnoses in 30% of the cases (n=56), three respiratory diagnoses in 7% (n=12), and none in 1% (n=3). No misclassifications were observed in the Severe Asthma Clinic program, whereas the AISBE program showed misclassifications of diagnoses in asthma (n=25) and bronchiectasis (n=40).

The WHO Disability Assessment Schedule (12-item WHODAS questionnaire)[45] reflected graded levels of disability in six domain-specific scores. The total WHODAS score was 1.88±0.77 (IQR 1.5– 2.4), consistent with light disability, as expected due to the entry criteria. Within the specific disability domains, a substantial interindividual dispersion was observed. Furthermore, a significant number of patients reported socioeconomic barriers, such limited financial resources (< 16k€/year; 74; 46.3%; 12.5% missing), or living alone (45; 24.6%), highlighting potential sources of inequity that could impact their self-management capabilities (**Appendices 2 and 3**).

### Design, components, and personalization of the Hybrid Care Intervention

#### Core and auxiliary components of the Hybrid Care Intervention

Throughout the co-design process, participants reached a consensus regarding the core elements of a patient-centered service that combines digital support with nurse-led care (**Figure 2**). With the following core elements: (1) nurse-led integrated care, (2) advanced and flexible digital support, and (3) patient-based health risk assessment. In addition, two auxiliary telemonitoring components: (4) A home-based monitoring set for the characterization and management of acute exacerbations; (5) IAQ assessment

**Figure 2.**
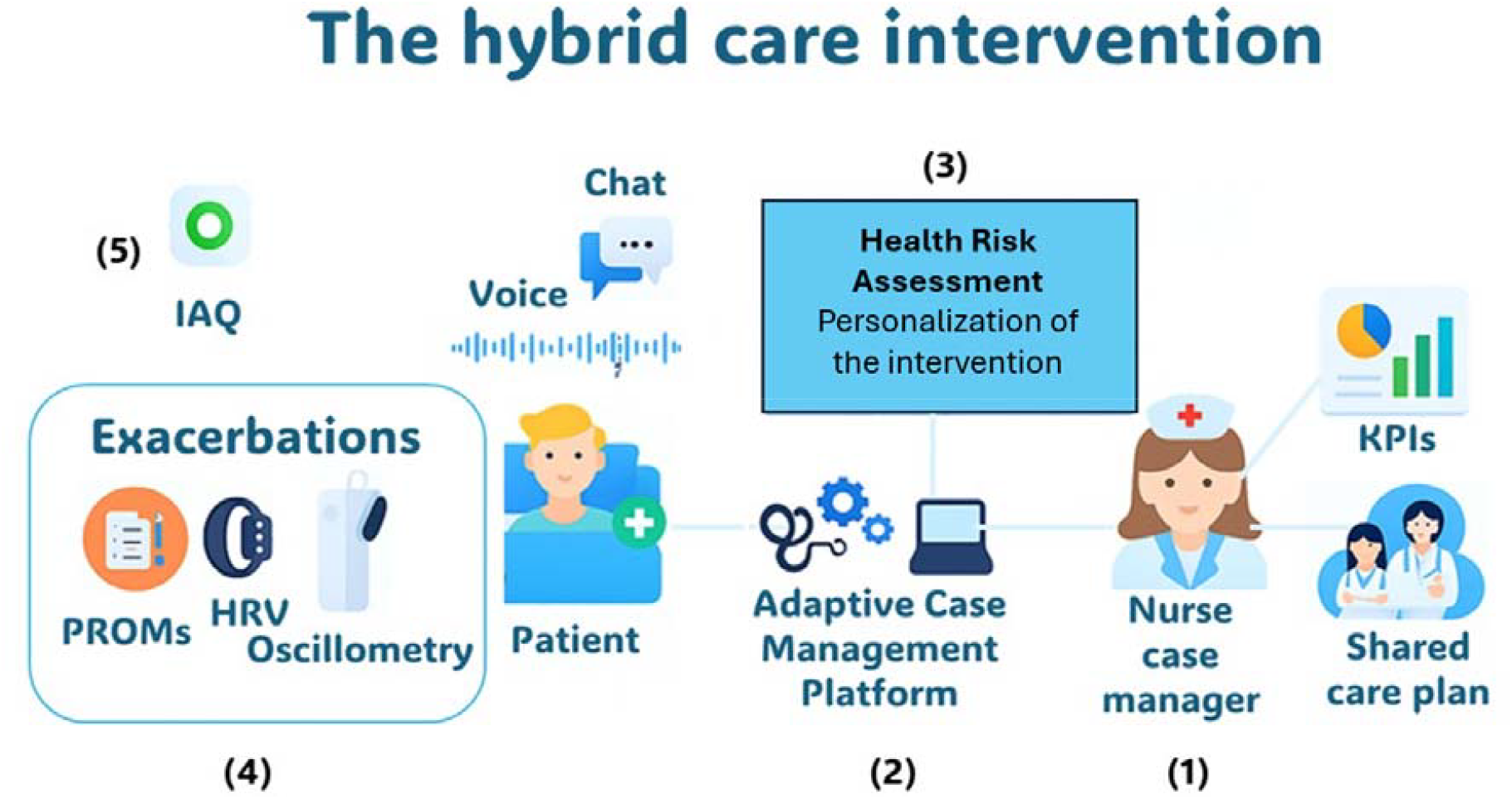
Components of the co-design process of the hybrid care intervention: (1) Roles of the nurse case manager, (2) Advanced Digital support using an Adaptive Case Management (ACM) Platform (Health Circuit)[18], (3) Personalization of the intervention through health risk assessment[46,47], (4) Characterization of acute episodes using daily home-based patient’s self-administered Oscillometry measurements (lung function testing)[48], heart rate variability (HRV) monitoring (autonomic regulation)[49], and symptoms assessed with a visual-analogic scale (VAS) modified from the CERT tool[50], and (5) assessment of household indoor air quality (IAQ) using low-cost sensors (LCS)[44]

#### The nurse case manager (Figure 2.1)

The management of clinically stable candidates begins with a holistic assessment followed by a personalization of the intensity and characteristics of the hybrid care intervention. The primary aim of the intervention is early management of exacerbations at community level preventing progression to severe acute episodes requiring emergency room visits and/or unplanned hospitalizations. It is acknowledged that the optimization of the approach requires patients’ proactivity and empowerment for self-management, ensuring communication with the nurse in an early phase of an acute episode. For the community-based program (i.e. AISBE), the nurse case manager is a trained professional administering the intervention tailored according to each patient’s profile. The reference primary care physician supervises the activity of the nurse, who also has easy access to specialized care (physicians and/or advanced care nurses) if needed. For specialized care programs (i.e. Severe Asthma Unit), the health professional leading the program has the profile of an advanced care nurse.

#### Digital support (Figure 2.2)

The digital support to the hybrid care intervention provided by the Health Circuit platform was reported in[18]. Briefly, the key features are: i) Multichannel communication between key professionals and patient ii) Shared care plans across healthcare tiers with an adaptive case management approach allowing flexibility in front of unexpected events and to specificities of different service providers, iii) Patients’ data capture (short questionnaires, testing and monitoring physiological data), iv) Cloud-based architecture to enable scalability and v) Standard-based interoperability with existing corporate-specific health information systems. To be highlighted that the digital tools provided to the patient are customized by the nurse case manager according to the patient’s needs at each time, taking into consideration their digital literacy.

#### Health Risk Assessment (Figure 2.3)

The HRA component represents the core analytical element of the hybrid care model, supporting patient profiling and the stratification of care intensity according to individual needs. It incorporates Machine Learning–based risk assessment models, currently under development, to provide: (1) identification of high-risk patients that would benefit the most from the hybrid care intervention, and personalization of the intervention guided by predictive modelling approaches, following the recommendations generated in[47]; and (2) decision support during exacerbations, by facilitating the objective identification of early signs of disease exacerbation to initiate preventive actions. In addition, the intervention includes transient home monitoring components implemented throughout the duration of acute episodes (1-2 weeks) for characterization and management of exacerbations.

#### Characterization of the exacerbations (Figure 2.4)

The pivotal aim is early identification and management of acute episodes based on objective grounds[20] through daily home-based patient’s data capture of Oscillometry (lung function testing), HRV (autonomic regulation) and symptoms’ assessment (VAS) during the exacerbations. The feasibility of the approach has been proven. Preliminary Oscillometry and VAS information is promising, and analysis of HRV is currently in process. Completion of characterization of exacerbations constitutes a central item within the fourth PDSA cycle.

#### Household IAQ assessment (Figure 2.5)

The recent report on continuous household IAQ monitoring using LCS done in the cohort[44] defines the role of IAQ assessment in selected cases. Available information does not support the need for household IAQ measurements on a routine basis.

### Clinical Management Pathways and Operational Features

Table 2 summarizes the clinical and operational characteristics of the hybrid care intervention, distinguishing between the community-based AISBE program and the specialized Severe Asthma Unit at HCB. The table details how the model articulates the roles of the main actors (patients, nurse case managers, and reference physicians) and the procedures for managing clinically stable and acute phases.

**Table 2.** Main roles and procedures associated with the hybrid care intervention.

| Elements in a Digitally Enabled Care Continuum Setting | Clinical programs |
| --- | --- |
| **Pro-active patient:** fulfilling inclusion criteria. Empowered for self-management. Accessibility to the nurse through a multimedia communication channel (*App*) and digitally enabled patient's self-capture data system which includes structured and non-structured data, including self-administered short questionnaires, and monitoring of physiological variables, such as HR, HRV, activity levels, pulse oximetry, etc..., using *wrist sensors*. | **Community-based AISBE program:** targeting co-morbid patients with chronic obstructive respiratory disorders and an AMG[26,27,31] scoring above percentile eighty in the health risk regional population pyramid (**Table 1**). The patients have as a reference the physician as the general practitioner. The interactions with other service modalities are fully aligned with the pre-existing shared care agreements at health district level (520 k citizens).<br>**Severe Asthma Clinic program:** targeting asthma patients, stages 5 and 6, controlled in the Severe Asthma Unit at HCB. One advanced care nurse could lead the program. |
| **Nurse case manager:** based in the community or at specialized clinics, depending upon the clinical program. Accessibility to the patient and to the reference physician (or other healthcare professionals) through a multimedia communication channel (*App*). Digitally enabled with the professional's dashboard |  |
| **Reference physician:** primary care physician or specialist depending upon the clinical program. Accessibility to the nurse case manager (to other |  |
| healthcare professionals and to the patient) through a multimedia communication channel (*App*). |  |
| **Management of clinically stable patients** |  |
| -The first step is the patient's motivational interview conducted by the nurse aimed at: (1) Empowering the patient for self-management of their condition, and (2) Facilitating the collaborative development of a patient's tailored care plan, with a holistic approach, agreed upon by the patient, the nurse, and the reference physician.<br>-The care plan forms the basis for the patient's periodic follow-up by the nurse case manager, with most interactions taking place through the established communication channel. In all cases, the management plan takes into consideration the diversity of patients' profiles.<br>-The clinical management plan is fully aligned with the standard care recommended for each respiratory diagnosis: COPD[1], Asthma[2] and Bronchiectasis[3]. |  |
| **Management of acute episodes** |  |
| -In the event of a worsening of clinical conditions, the patient can contact the nurse case manager through the communication channel. The contact triggers the process of identifying a potential exacerbation, based on the patient's reported symptoms (mMRC and CAT), clinical judgement of the nurse, including Oscillometry measurement. The nurse will be able to interact with the reference physician to decide on further steps needed for the diagnosis and management of the acute episode due to i) lung-related events, ii) non-pulmonary dyspnea, or iii) dyspnea of mixed origin.<br>- Home-based management of the acute episode activates: i) close follow-up of the patient, remote or face-to-face, by the nurse case manager, ii) daily patient's self-administration of short questionnaires (VAS), as well as measurements of Oscillometry and monitoring of physiological variables (HRV and activity) with a wrist device, and iii) adjustment of the therapeutic plan in agreement with the reference physician.<br>This action plan will be maintained for one or two weeks depending upon the patient's progress. |  |

*HR: Heart Rate; HRV: Heart Rate Variability; AISBE: Integrated Social and Health Care Based on Evidence; AMG: Adjusted Morbidity Groups; HCB: Hospital Clínic de Barcelona; HRV: Heart Rate Variability; mMRC: Modified Medical Research Council Dyspnoea Scale; CAT: COPD Assessment Test; COPD: Chronic Obstructive Pulmonary Disease; VAS: Visual Analogic Scales.*

### Tailoring of the intervention: Patient profiles and digital competencies

Personalization of the hybrid care intervention emerged as a key outcome of the co-design process Tailoring was approached from three complementary perspectives: (1) clinical profiling of candidates, (2) contextual factors modulating care (socio-economic, disability, living conditions, etc…); and (3) assessment of digital literacy to match the level of technological support to individual capabilities and needs.

#### Candidate profiling

The baseline assessment of the patients and the contacts between the nurse case manager and participants over the first 18 months of the follow-up enabled the identification of six clinical groups: two within the Severe Asthma program (A1–A2) and four within the community-based AISBE program (C1–C4) (**Appendix 1**). These profiles formed the basis for personalizing the intensity and nature of the hybrid care components outlined in **Table 2**. The groups and their corresponding hybrid care implications are detailed in **Table 3**. Assessment, and further refinement, of the profiled hybrid care intervention for five (A1, A2 and C2-C4) out of the six patients’ groups (**Table 3**) will be consolidated during the fourth PDSA cycle. Further information on key baseline characteristics across clinical groups is displayed in **Appendices 2-3**.

**TABLE 3.** The six clinical groups defined in the study cohort.

| Grup | Main features | Hybrid Care implications |
| --- | --- | --- |
| **Severe Asthma Program at HCB (n=48)** |  |  |
| **A1**<br>23 (47.92%) | Severe asthma, stages 5 and 6[28,29], clinically stable without requiring interactions with the nurse | Provide access to the communication channel (*App*) without scheduling specific actions with the nurse. |
| **A2**<br>25 (52.08%) | Severe asthma, stages 5 and 6 and exacerbations and/or other problems (mental disorders: mostly anxiety/depression, social frailty, ...) trigger frequent interactions with the nurse | Target candidates for a hybrid care intervention. Management by an advanced care respiratory nurse. Specific needs were identified for A2 patients with severe asthma and obesity. |
| **Community-based AISBE Program (n=135)** |  |  |
| **C1**<br>50 (37.04%) | Mild-moderate lung disease (i.e. COPD, GOLD I or II) with few symptoms and unproblematic co-morbidities. | Not candidates for the intervention. |
| **C2**<br>18 (13.33%) | Moderate-severe lung disease, and/or rapid disease progress and/or history of severe exacerbations | Target candidate for the hybrid care intervention with shared care agreements with specialized care |
| **C3**<br>33 (24.44%) | Moderate-severe lung disease with problematic co-morbidities and/or social frailty. | Target candidate requiring personalized medical (co-morbidities) and social support [47] |
| **C4**<br>34 (25.19%) | Very severe lung disease (i.e. COPD, GOLD IV) is often under home respiratory therapies. | Target candidate interacting closely with specialized care through an advanced care respiratory nurse. |

#### Digital literacy

The study evaluated the participants’ knowledge, usage, and confidence across a range of digital skills relevant to the hybrid care digital solution. Briefly, 109 patients showed either high digital skills (n=51; 27.9%) or medium level scoring with significant improvements after training (n=58; 31.7%). The remaining patients presented significant limitations in the use of digital tools ranging from: i) unable (n=16; 8.7%), ii) poor digital skills (n=37; 20.2%), and iii) medium level scoring without significant improvements with training (21; 11.5%) (**Appendix 4**). Overall, 41 (22.4%) participants did not use the app, either per the nurse’s evaluation or by the participant’s own decision. The two most critical functions limiting the use of digital tools were: i) login to the App, and ii) synchronization of the wrist sensor.

The co-design process confirmed feasibility and stakeholders’ acceptance of the overall approach. Likewise, it contributed to identify some barriers partly limiting the digitally enabled management of the cohort and provided emergent solutions for those limitations (**Table 4**). These refinements will be addressed in future stages of the project, which will take place in a final PDSA-cycle to be completed between September 2025 and end of February 2026.

**Table 4.** Expected refinements of the hybrid care intervention during the fourth PDSA cycle.

| Intervention Components | Expected Improvements & Outcomes |
| --- | --- |
| **Digital support** | o Easy patient two-factor authentication with biometrics (e.g., biometric two-factor authentication) to address login barriers<br>o Reduce the time required for synchronization of the wearable clinical device for monitoring of Heart Rate Variability<br>o Expand multimedia communication channel to other professionals<br>o Enhanced clinician's dashboard<br>o Design of a novel voice-optimized interface, based on conversational agents powered by generative artificial intelligence (GenAI) |
| **Oscillometry** | o Develop and validate clear guidelines for home-based Oscillometry use during exacerbations<br>o Create data-driven parameters for the management of moderate exacerbations at the community level. |
| **Foster sustainable adoption[51,52]** | o Refine personalized intervention protocols for the six identified clinical profiles (Table 3).<br>o Expand engagement of all healthcare professionals<br>o Deployment of training programs for professionals<br>o Identification of KPIs facilitating program management<br>o Booklet guiding the implementation process<br>o Assessment of healthcare value generation[53] (*) |
| **Risk assessment & Clinical Decision Support** | o Identification of high-risk candidates<br>o Decision support tools for personalization of the intervention<br>o Parametrization of management of moderate exacerbations<br>o Proof of concept of the potential role of GenAI tools |

(\*) *Quintuple Aim Assessment*: (1) health outcomes, (2) patient reported outcomes (PROMs) and patient reported experiences (PREMs), (3) health professionals' experience, (4) direct costs evaluated with a cost-consequence analysis and (5) estimation of indirect costs to assess potential sources of inequity.

Several limitations in clinical information quality and continuity emerged as major barriers to implementation: (1) diagnostic inconsistency in patients’ electronic care records, particularly in asthma and bronchiectasis, highlighted the need for stronger alignment between coded diagnoses and validated clinical criteria; (2) In COPD, incomplete documentation of disease trajectories and progression markers prevents the establishment of longitudinal baselines, which are essential for timely risk prediction and adaptive management; (3) functional and disability assessments, especially those capturing cognitive, psychosocial, and social determinants, remain largely absent from electronic health records, leading to an incomplete view of patient vulnerability.

#### Implementation of the hybrid care intervention beyond PDSA-4

Within the co-design process several well-known consolidated frameworks guiding implementation processes were evaluated aiming at fostering clinical adoption of the hybrid care intervention. A pragmatic approach to the Integrated Theory-based Framework for Intervention Tailoring Strategies (ItFits-Toolkit)[51,52] was proposed to provide guidance. The two key features triggering the selection of these two implementation Toolkits were: (1) high potential for a pragmatic application in the real-world setting covering the entire implementation cycle, and (2) plans for future use of these implementation tools at regional level.

## DISCUSSION

The study highlighted the feasibility and acceptance of a hybrid care intervention integrating nurse-led management, digital support, and patient-based health risk assessment for high-risk respiratory patients. The current proposal is grounded on evidence indicating that these three components of the hybrid care intervention can significantly decrease the burden of chronic obstructive respiratory disorders by reducing potentially avoidable hospitalizations which should enhance patients’ quality of life and prognosis while reducing health care costs [12,13,17,18]. However, the study clearly indicates the need for addressing the practicalities of this still novel approach through a co-design process with participation of the key stakeholders. Tailored implementation[51,52] and assessment of value-generation[53] in the real-world should be the next steps before scalability as routine care.

Recent reports indicate that prevention, and enhanced management, of community-based exacerbations in patients with COPD should be a priority that may improve patient trajectories and outcomes[4]. In this scenario, nurse-led personalized interventions with digital support have been proposed as a novel modality of care aiming at reducing potentially avoidable hospitalizations[5]. Moreover, the approach involves opportunities for enhanced patient stratification[46] contributing to disentangling mechanisms of obstructive respiratory disorders, rethink disease taxonomies and, ultimately, open new avenues for clinical research in these patients.

From the operational point of view, the co-design process allowed the identification of structural and operational gaps in current practice that hinder the full deployment of the hybrid care intervention. Therefore, these limitations must guide the prioritization of forthcoming refinements.

Finally, digital literacy emerged as a critical determinant of patients’ engagement. From an implementation perspective, these findings highlight a dual challenge: (1) Ensuring that fragile and socially vulnerable patients are not excluded from digitally enabled care, and (2) Allocating resources for tailored training, simplified technological solutions, and/or caregiver-mediated support. Our findings reinforce the strategic role of the nurse case manager, not only delivering clinical care, but also acting as a main facilitator of digital adoption.

Therefore, we acknowledge that further refinements of the hybrid care proposal are still required before its deployment in routine clinical care. These refinements will be addressed in a final PDSA-cycle to be completed between September 2025 and end of February 2026.

## CONCLUSIONS

The current study indicates that the adaptive case management approach for personalized hybrid care among high-risk chronic obstructive respiratory patients at the community level is feasible and well accepted by the different stakeholders. Through a sequence of PDSA cycles, we confirmed the need for tailored implementation before full integration into day-to-day practice and subsequent scaling at the system level. The study outputs provide guidance on professional roles and patient profiling to support consensus-based adaptation of the intervention. Future PDSA cycles will refine this guidance by identifying key elements to enhance implementation fidelity and promote equity in healthcare delivery.

## Abbreviations

ACM: Adaptive Case Management
AISBE: Integrated Health District of Barcelona-Esquerra
AMG: Adjusted Morbidity Groups
CAT: COPD Assessment Test
CHSS: Catalan Health Surveillance System
COPD: Chronic Obstructive Pulmonary Disease
EU: European Union
FEV_1_: Forced Expiratory Volume in 1 second
FEV_1_/FVC: Ratio of FEV_1_ to FVC
FVC: Forced Vital Capacity;
HCB: Hospital Clínic de Barcelona
HR: Heart Rate
HRV: Heart Rate Variability
IAQ: Indoor Air Quality; IT: Information Technology
KPI: Key Performance Indicator
LCS: Low-Cost Sensors
mMRC: Modified Medical Research Council Dyspnoea Scale
PCA: Principal Component Analysis
PDSA: Plan-Do-Study-Act
PREMs: Patient Reported Experiences Measures
PROMs: Patient Reported Outcomes Measures
RCT: Randomized Controlled Trial
SD: Standard Deviation.

## Data Availability

The datasets generated and/or analysed during the current study contain sensitive patient information and will not be openly distributed. However, anonymized data may be made available upon reasonable request to the corresponding author, subject to institutional data sharing agreements and ethical approval.

## Authors’ Contributions

NS and EA were the clinical leads of the project, with the support of all members of the clinical team; NSR, MS, MP, JSY, and MJG. RGC and JR coordinated the local implementation of the project within the framework of the K-HEALTHinAIR initiative, under the consortium leadership of AA and JF. AGL was responsible for patient recruitment and follow-up of the study cohort. The digital tools were provided by Health Circuit under the supervision of IC. RF contributed with expert advice on the use of Oscillometry. EV and RGC prepared the databases and performed the statistical analyses and the figures. In addition, AGL, NSR, MS, RGC, MP, JSY, MJG, GCS, AJ, XM, JR, IC, NS, and EA participated in the focus group activities that supported the co-design process of the intervention. The first draft of the manuscript was jointly written by AGL, RGC, IC, JR, EA, and NS. All listed authors contributed to the review and editing of the final version of the manuscript and approved its submission.

## Acknowledgements

The K-HEALTHinAIR project funded this study, Grant Agreement nº 101057693, under a European Union’s Call on Environment and Health (HORIZON-HLTH-2021-ENVHLTH-02).

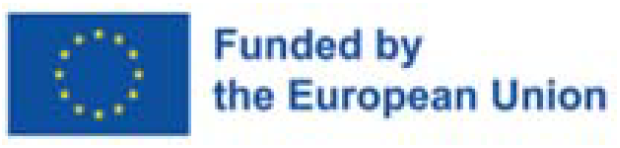

## Disclaimer

Views and opinions expressed are, however, those of the authors only and do not necessarily reflect those of the European Union or the European Health and Digital Executive Agency as granting authority. Neither the European Union nor the granting authority can be held responsible.

