## Supplementary material for "A hybrid care intervention for high-risk patients with chronic obstructive respiratory diseases: bridging the gap between clinical trials and real-world practice"

*Alba Gómez-López et al.*

*-*

**Supplementary Material**

Contents

Appendix 1. The [co-design process	2](#_Toc211977578)

Appendix 2. Baseline characteristics across clinical groups	[3](#_Toc211977579)

Appendix 3. Disability assessment across clinical groups	[4](#_Toc211977580)

Appendix 4. [Digital literacy assessment	5](#_Toc211977581)

### APPENDIX 1 – The co-design process

The co-design process was structured into four sequential Plan–Do–Study–Act (PDSA) cycles conducted over a two-year period, from January 2024 to December 2025, as outlined in the study protocol[1]. The originally planned six-month duration of each PDSA cycle was slightly modified, with a two-month extension of PDSA-3 to enable full maturation of the core characteristics of the hybrid care intervention. The positive acceptance of the approach by the various stakeholder groups prompted the preparation of this report and the initiation of the fourth PDSA cycle, which runs from September 2025 to February 2026 (**Table S1 and Table 4**). Subsequent steps will include the tailored implementation of the hybrid care intervention in real-world settings beyond the current study cohort, as well as an evaluation of its healthcare value generation.

During the first three PDSA cycles, the four members of the core research team: nurse case manager (AG), a respiratory specialist (JR), a data scientist (RG), and a digital health specialist (IC) jointly refined the different components of the hybrid care intervention (**Figure 2**) through continuous interaction with participants in the study cohort (n=205). The hybrid care intervention was implemented across two clinical programs: (i) integrated care for AISBE (asthma, COPD, and bronchiectasis), jointly led by respiratory specialists (NS) and primary care professionals (NS), representing 80% of the study cohort; and (ii) the Severe Asthma Unit at HCB, led by EA, encompassing the remaining 20% of participants. These clinical leaders provided sustained guidance and feedback throughout the co-design process. In addition, partners from the EU project *K-HealthinAir* (JF and AA) contributed continuous input on technical aspects of household indoor air quality (IAQ) monitoring, while RF offered expertise on Oscillometry-related topics. Several workshops and meetings with the collaborators mentioned above were held through the co-design process.

At the conclusion of PDSA-3, an early draft of the main manuscript was shared with the participants in a two-hour focus group session held in early September 2025. The session addressed three key questions: (i) the structure and components of the hybrid care intervention; (ii) barriers and facilitators for its tailored implementation in real-world settings; and (iii) the clarity and coherence of the draft manuscript. The focus group provided valuable insights, and all participants contributed to refining the final version of this manuscript.

The fourteen focus group participants represented a diverse range of professional profiles, including respiratory specialists (EA, MP, NS, JSY, JR), primary care professionals (NS, MJG, MS, AG), digital health specialists and data scientists (IC, XM, RG), and healthcare managers (GC, AJ).

**TABLE S1 – Adapted co-design protocol for the enhanced management of multimorbid patients with COPD or severe asthma: role of IAQ**[1].

| **Analysis of the relationships between IAQ and health status in the patients’ cohort** **(n=205) within the EU project K-Health in Air** (<https://k-healthinair.eu/>), **and tailored implementation plans for sustainable adoption of the hybrid care intervention in real-world settings** | |
| --- | --- |
| **# Cycle** | **Objectives** |
| **PDSA-1**  *(January – June 2024)* | Analysis of feasibility of the four components of the hybrid care intervention as planned in the seminal protocol^22^. |
| **PDSA-2**  *(July – December 2024)* | Finetuning of the four core components of the intervention following the requirements identified in PDSA-1. |
| **PDSA-3 (*)**  *(January – August 2025)* | Completion of finetuning of the components of the intervention |
| **PDSA-4**  *(September 2025 – February 2026)* | (1) Progress towards adoption of the basic version of the hybrid care intervention; (2) Evolutions of the components of the intervention |

*^(*)^ Timeframe of the current report, from PDSA-1 to PDSA-3; IAQ: Indoor Air Quality; LCS: Low-Cost Sensors; Symptoms:  mMRC/CAT questionnaires; HR: Heart Rate; HRV: Heart Rate Variability; PDSA: six-month co-design cycles using Plan-Do-Study-Act[2] methodology.*

### APPENDIX 2 - Disability assessment across clinical groups

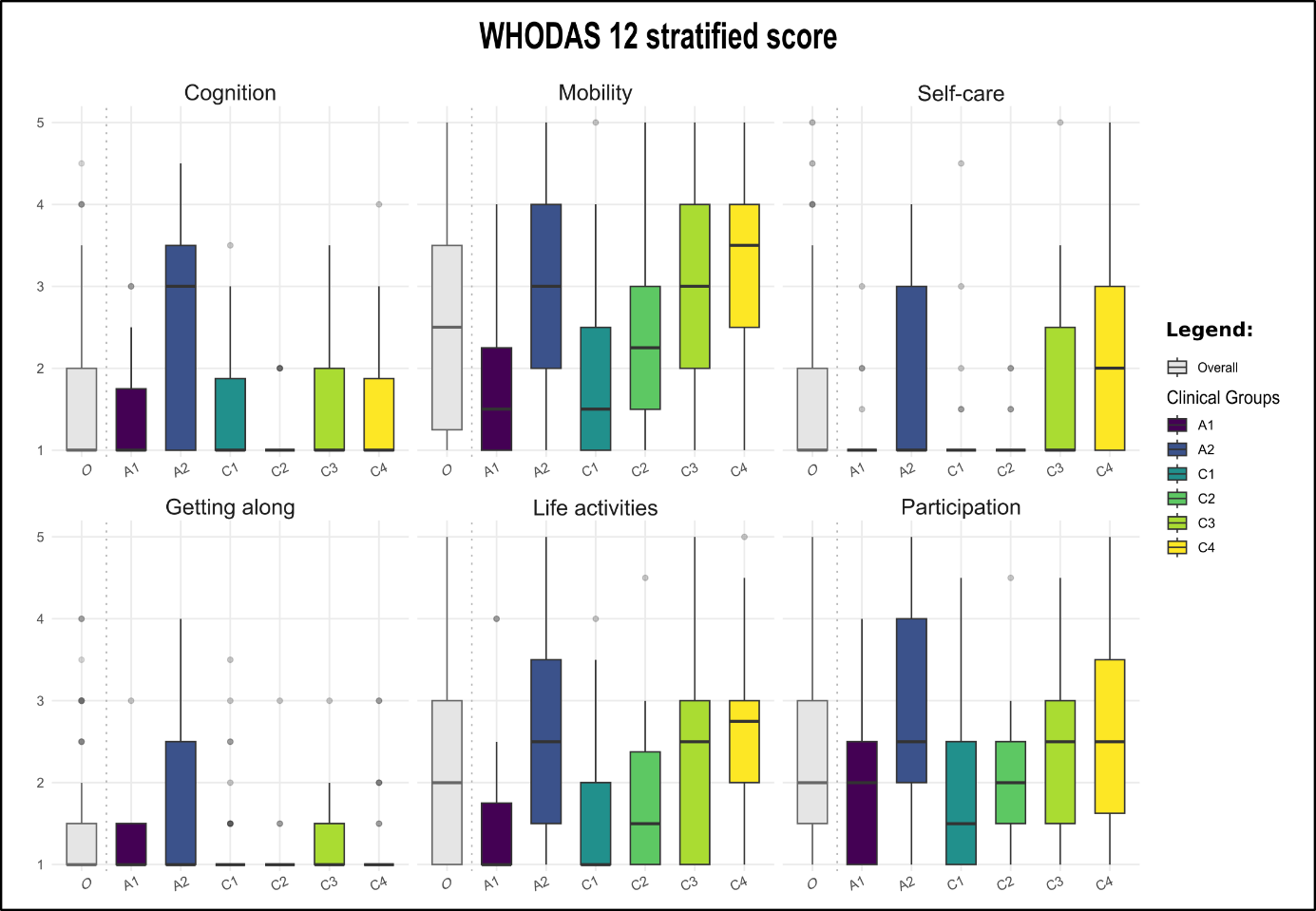

***Figure S1 – Disability assessment*** *using the WHODAS 12.0[3] questionnaire. Boxplots of the assessment of disability for the six dimensions of the questionnaire. The first boxplot corresponds to the entire study cohort, whereas the subsequent six boxplots correspond to the results obtained for each of the clinical groups. Disability domains: Cognition (understanding/communicating), Mobility (moving/getting around), Self-care (hygiene, dressing, eating, staying alone), Getting along (interacting with others), Life activities (household/work), and Participation (community participation and societal involvement).*

**Clinical Groups Overview**

**A1:** clinically stable severe asthma (steps 5–6), infrequent nurse interactions.
**A2:** unstable severe asthma with psychosocial comorbidity (e.g., anxiety/depression, social frailty); frequent nurse interactions.
**C1:** COPD/bronchiectasis with mild–moderate impairment, few symptoms, non-problematic multimorbidity.
**C2:** moderate–severe respiratory disease and/or rapid progression and/or prior severe exacerbations.
**C3:** moderate–severe respiratory disease plus problematic multimorbidity and/or social frailty.
**C4:** very severe disease (often COPD GOLD IV), frequently on home respiratory therapies.

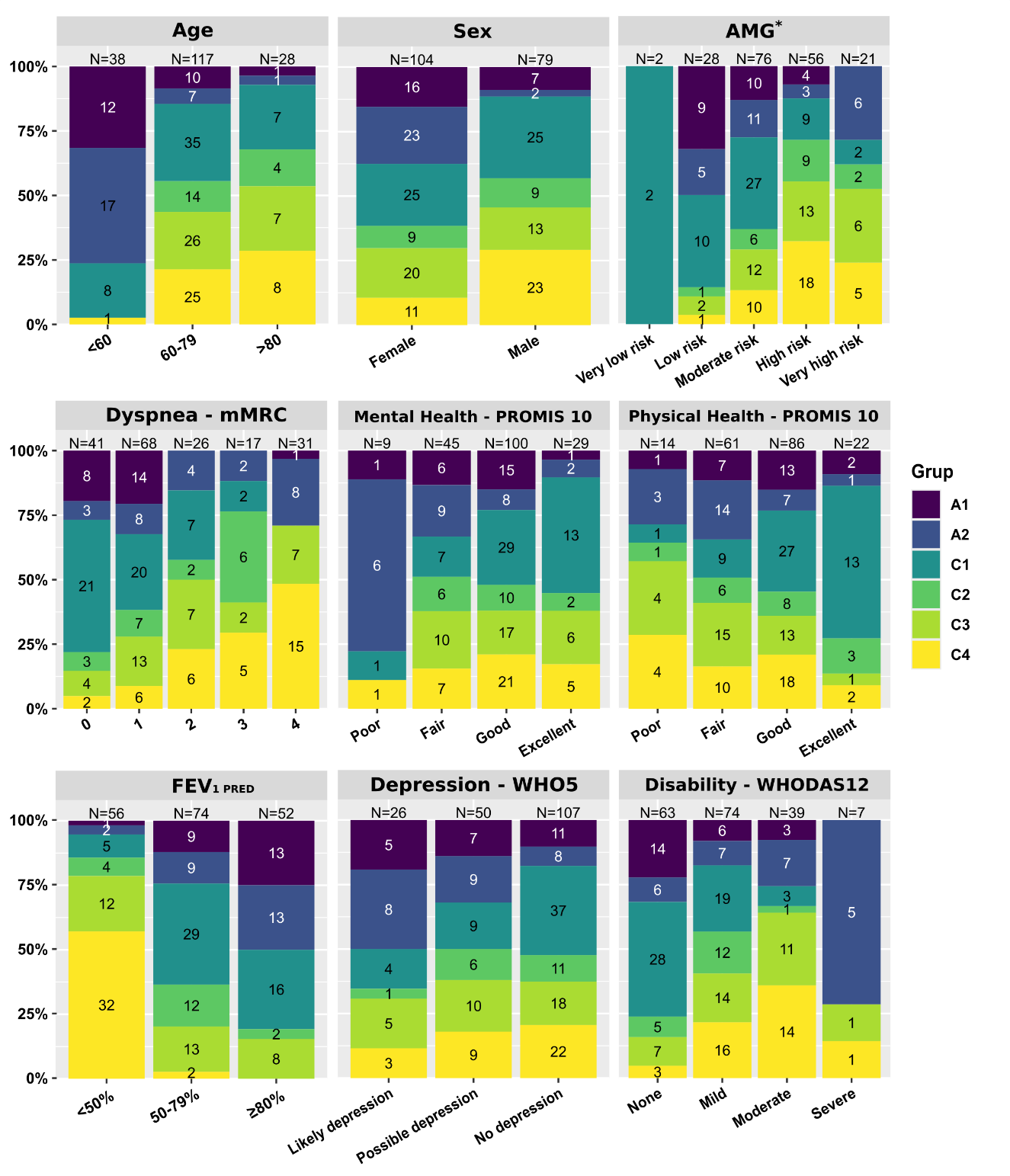

### APPENDIX 3 - Baseline characteristics across clinical groups

***Figure S2. Stacked distributions of key baseline characteristics across clinical groups.*** *Stacked bar charts showing the relative distribution of the six clinical groups (A1–A2: Severe Asthma program; C1–C4: AISBE community program) across baseline variables. Panels depict age, sex, AMG[4,5] risk strata, WHO-5 well-being[6], PROMIS-10 Global Mental and Physical Health[7], airflow-limitation, Dyspnoea[8], and WHODAS-12[3] disability severity. Bars are normalized to 100% within each category; numbers inside bars are counts; and numbers above facet strips indicates the number of observations for that level. * AMG: Adjusted Morbidity Groups score.*

### APPENDIX 4 - Digital literacy assessment

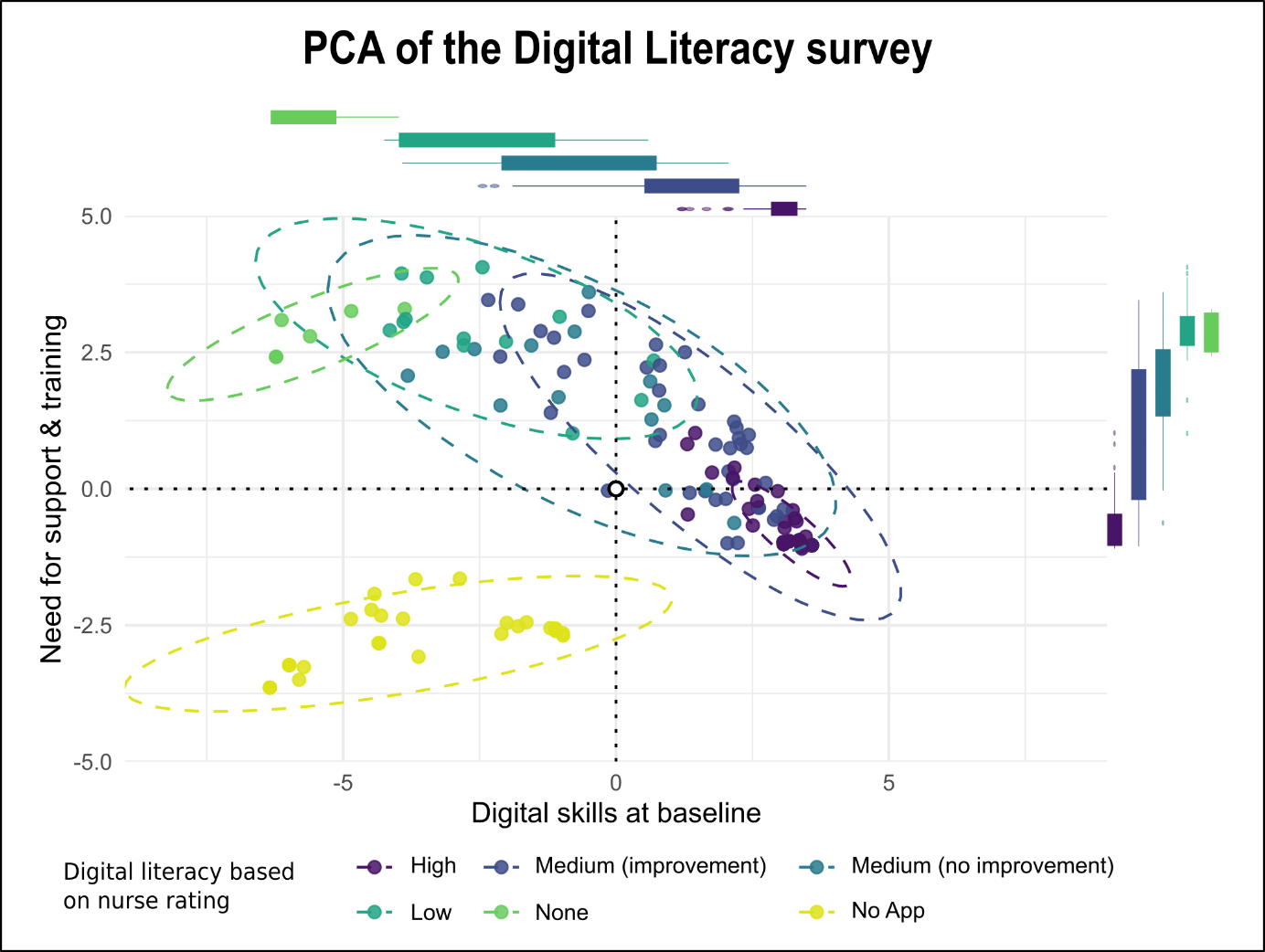

***Figure S3: Digital literacy.*** *Distribution of the patients (dots) according to their baseline digital literacy scoring (X-axis) against the requirements in terms of support and training (Y-axis) resulting from the twelve questions evaluated in the survey.* Categorical responses were binarized, and a Principal Component Analysis (PCA) was applied to identify underlying patterns. *The colors indicate the six groups resulting from the digital literacy scoring subjectively assessed by the nurse case manager: i) high digital skills (garnet); ii) medium scoring with improvement after training (dark blue), and iii) without improvement after training (dark green); iv) low digital skills (lighter green); v) no skills after using the digital tools (light green); and vi) digitally excluded by the nurse, never used the digital tools available in the study (yellow. The discontinuous lines indicate the area including 95% of the individuals in each of the six groups identified). The boxplots of digital skills (X-axis) and requirements of support (Y-axis) for the five groups using the digital tools are also depicted in the figure.*

**TABLE S2 – Digital literacy by clinical program**

| **Digital literacy** | **All patients (n=183)** | **Patients followed by Primary Care (n=135)** | **Patients followed by the Severe Asthma Unit (n=48)** | **p- value** |
| --- | --- | --- | --- | --- |
| None; n (%) | 16 (8.74%) | 15 (11.11%) | 1 (2.08%) | .012 |
| Low; n (%) | 37 (20.22%) | 30 (22.22%) | 7 (14.58%) |  |
| Medium (no improvement); n (%) | 21 (11.48%) | 18 (13.33%) | 3 (6.25%) |  |
| Medium (can improve); n (%) | 58 (31.69%) | 43 (31.85%) | 15 (31.25%) |  |
| High; n (%) | 51 (27.87%) | 29 (21.48%) | 22 (45.83%) |  |
